# Measuring Icebergs: Using Different Methods to Estimate the Number of COVID-19 Cases in Portugal and Spain

**DOI:** 10.1101/2020.04.20.20073056

**Authors:** Carlos Baquero, Paolo Casari, Antonio Fernández Anta, Davide Frey, Augusto Garcia-Agundez, Chryssis Georgiou, Raquel Menezes, Nicolas Nicolaou, Oluwasegun Ojo, Paul Patras

**Affiliations:** U. Minho & INESC TEC, Portugal; University of Trento, Italy; IMDEA Networks Institute, Spain; Inria Rennes, France; TU Darmstadt, Germany; U. Cyprus, Cyprus; U. Minho, Portugal; Algolysis Ltd, Cyprus; IMDEA Networks & UC3M, Spain; U. Edinburgh, UK

## Abstract

The world is suffering from a pandemic called COVID-19, caused by the SARS-CoV-2 virus. The different national governments have problems evaluating the reach of the epidemic, having limited resources and tests at their disposal. Hence, any means to evaluate the number of persons with symptoms compatible with COVID-19 with reasonable level of accuracy is useful. In this paper we present the initial results of the @CoronaSurveys project. The objective of this project is the collection and publication of data concerning the number of people that show symptoms compatible with COVID-19 in different countries using open anonymous surveys. While this data may be biased, we conjecture that it is still useful to estimate the number of infected persons with the COVID-19 virus at a given point in time in these countries, and the evolution of this number over time. We show here the initial results of the @CoronaSurveys project in Spain and Portugal.

## 1 Introduction

During the current coronavirus pandemic, monitoring the evolution of COVID-19 cases is very important for the authorities to make informed policy decisions, and for the general public that has the right to be informed of the reach of the problem. Official numbers of confirmed cases are periodically issued by each country’s health authority. (For Portugal see [1].) Unfortunately, upon the pandemic outbreak is it usually the case that there is lack of available laboratory tests, and other material and human resources. Hence, it is not possible to test all potential cases, and some eligibility criteria is applied to decide who is tested. Under these circumstances, the evolution of official confirmed cases might not represent the total number of cases (see [5]).

This motivates the need of other probing techniques, beyond laboratory testing, that may bring in information about the potential numbers of cases. In fact, it is very likely that the true numbers might only be known in the future, once serological surveys are done (as happened in prior outbreaks [10]). However, any rough estimate that is of the same order of magnitude as the real number will be better than being almost in the dark.

In the rest of this document we present the @CoronaSurvey project [7], which uses a new approach to estimate the number of cases with COVID-19 symptoms, based on using crowdsourcing via open anonymous surveys to obtain indirect information. We compare this new approach with approaches that infer current cases from the case fatality series for two particular countries, Portugal and Spain.

## 2 Delay-adjusted Case Fatality Ratio

If we consider the current number of fatalities and divide it by the current number of confirmed cases, we obtain a naive case fatality ratio (CFR). However, this metric is not using the actual number of cases with known outcomes, since recent cases will still evolve into fatalities and recoveries. By estimating the true number of cases with known outcomes, it is possible to obtain a corrected case fatality ratio (cCFR) that takes into account the average delay from symptoms to death, as in [6]. Since the corrected denominator (cases with known outcomes) is reduced, the cCFR is higher than the naive CFR during a growing outbreak. A high cCFR is typically an indicator of lack of coverage in laboratory testing. If we assume, in general, that the disease will have similar case fatality rates in different countries, it is possible to use the known fatality ratio from Wuhan, China, (currently at 1.38%). This value is compared with the obtained cCFR, and the percentage of coverage (the proportion of cases that are, in fact, confirmed) in different countries can be estimated. This is done in [8] and, for instance, the projected coverage in Portugal and Spain is 19% and 4.7%, respectively. These values can in turn be used to correct the number of reported confirmed cases in each country, and estimate the likely true number of cases, thus probing the iceberg that lurks under water.

Another technique, based on the same corrections for delay, but using an overall gross value for estimating the true cases from the mortality rate is simply obtained by multiplying the cumulative mortality by 400. See [5]. We will use both techniques for estimation in each country.

## 3 Crowdsourcing with Open Anonymous Surveys

The @CoronaSurveys effort is based on crowdsourcing data collection, in a way that avoids querying the citizens about their particular health status and identity. Participants have to fill a simple survey with three questions (we currently use Google Forms). In the first question participants select a geographical area, which can be a whole country or a region within a country. Then, they answer two simple questions:

- How many people do you know in this geographical area? (Please, consider only people whose current health status you likely know.)
- As far as you know, how many of the above have had symptoms compatible with COVID-19 (or were diagnosed with the disease)?

By not asking any personal information we aim at protecting the participant’s privacy, and having just two questions aims to simplify the answering process. This will hopefully increase the participation. However, the lack of detailed information of the participants makes the estimation process challenging. We do not control the spread of the survey nor the adequate coverage of regions, age groups and other parameters. Somewhat surprisingly, even given these limitations, we can still obtain a rough estimate and see that it is not far from those obtained with other techniques. One reason for this may be that each participant is typically reporting on the health status of a large sample (hundreds), which increases significantly the coverage of the survey. Obvious advantages of this approach are that it is very simple to deploy and can give very timely results.

The process to obtain the estimate of cases is as follows. Survey responses are cleaned by identifying and removing outliers. These are answers unusually large in terms of the range of persons that the respondent declares to know (we remove entries outside 1.5 times the interquartile range above the upper quartile), and the ratio of symptomatic people reported (we remove entries above 30% of reporting). The latter are removed because we aim at surveying the general population that is not in particular high contact with symptomatic cases. Once the data is clean, we use it to obtain the percentage of cases in the population that is known to the respondents. Then, we naively extrapolate this ratio to the whole country population.

Our initial surveys were done on Twitter and did not ask for how many people are known to the respondents. For those, the number of known people was set to 150, which is the Dunbar number [9, 2], the expected number of persons with whom one maintains stable social relationships.

In the following sections we show the results we obtain for the two countries, Portugal and Spain.

## 4 Estimating the Number of COVID-19 Cases in Portugal

Portuguese data on fatality and confirmed cases was obtained from [4], a public repository that helps disseminate the official DGS daily data.

In Figure 1 a comparison of the number of cases accounted for or estimated with the described techniques is presented. The solid line plots the number of officially reported confirmed cases. The dashed line is obtained by multiplying the number of fatalities by 400, while the solid dots • indicate the more precise estimate from the corrected case fatality rates, and compensating for the estimated coverage of testing. Finally, the diamond ⋄ represents our estimates from the two initial Twitter surveys, and the Δ triangles the results from the subsequent two more complete open surveys.

**Figure 1:**
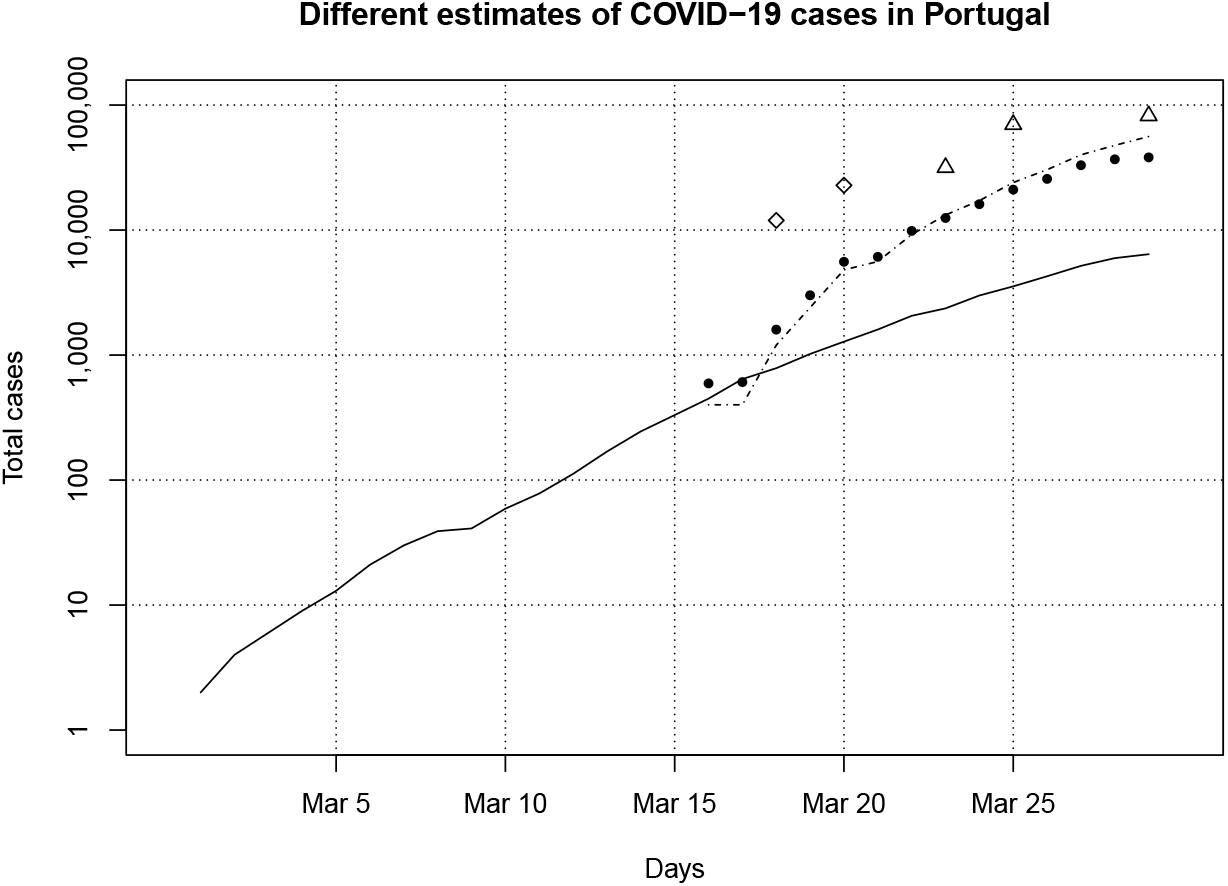
Case estimates for Portugal, March 29, 2020. Solid line is official confirmed cases. Dashed lines and solid dots • are the number of cases as estimated from fatalities (times 400) and from corrected case fatality ratios, respectively. Diamonds ⋄ and triangles Δ show the results from @CoronaSurveys, Twitter and Google Forms, respectively.

As we can see, the anonymous surveys, by @CoronaSurveys, tend to estimate more cases, likely over-estimating. In any case, these estimates still follow the overall tendency, and seem to be closer to the order of magnitude of the true numbers than the official reports.

## 5 Estimating the Number of COVID-19 Cases in Spain

Spanish data was obtained from the European Center for Disease Control [3], which aggregates public information about COVID-19 cases across the World.

For the Spanish case (see Figure 2) there is an even wider gap between the number of cases predicted from fatality numbers and the reported cases. For the estimates based on open surveys, we include data points from one Twitter survey and three subsequent Google Forms surveys as described in the previous section. Surprisingly, the results from @CoronaSurveys are very close to the fatality-based estimators, while providing still slightly higher estimates.

**Figure 2:**
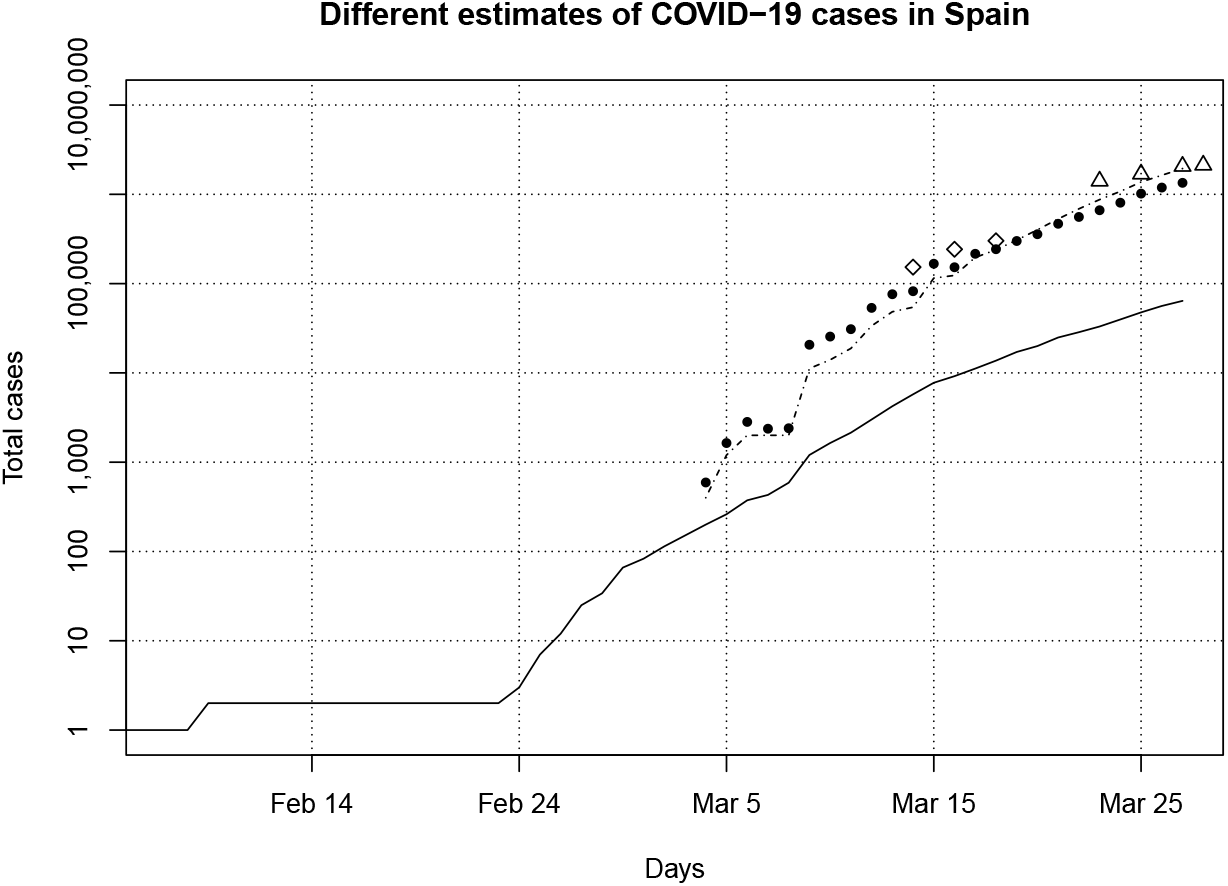
Case estimates for Spain, March 29, 2020. Solid line is official confirmed cases. Dashed lines and solid dots • are the number of cases as estimated from fatalities (times 400) and from corrected case fatality ratios, respectively. Diamonds ⋄ and triangles Δ show the results from @CoronaSurveys, Twitter and Google Forms, respectively.

## 6 Discussion

In this first report we draw attention to the limitations of relying only on confirmed cases to measure the true size of a growing pandemic. From existing data it is possible to derive other measures, and, surprisingly, it is also possible to do simple surveying approaches that, while simple and maybe crude, are clearly non invasive of privacy, and still get meaningful data. In particular @CoronaSurveys can be relevant in countries with a decent digital infrastructure but lacking in laboratory resources. When measuring icebergs, there are many strategies.

## Data Availability

All the data used is openly available.

https://github.com/GCGImdea/coronasurveys/tree/master/data

## 7 Further Progress and Acknowledgments

As mentioned, this document is the first report on the @CoronaSurveys project [7], describing its status at the end of March 2020. Since then, the project has grown rapidly, with specific surveys open for 24 different countries in 13 different languages, plus two generic surveys open for the rest of countries in English and Arabic. All the data collected and several estimates computed are openly available at the project web site [7], which also contains plots of the estimates for all countries.

We would like to thank the large group of resaerchers and collaborators that is currently involved in the @CoronaSurveys project in any form: Payman Arabshahi, Annette Bieniusa, Elisa Cabana, Ignacio Castro, Angeliki Gazi, Benjamin Girault, Mathieu Goessens, Harold Hernández, Rodrigo Irarrazaval, Anna Ishchenko, Oleksiy Kebkal, Rosa Lillo, Alvaro Méndez, Esteban Moro, Antonio Ortega, Yuichi Tanaka, Christopher Thraves, Erol Sahin, Andres Schafer, Ghadi Sebaali, Natalie Soto, Matias Spatz Fernández, Efstathios Stavrakis, Pelayo Vallina, and Lin Wang.

## References

[1] Portugal Direção Geral da Saúde, Ministério da Saúde. COVID-19. https://covid19.min-saude.pt. Accessed: 2020-04-09.

[2] Robin Dunbar. How many friends does one person need?: Dunbar’s number and other evolutionary quirks. Faber & Faber, 2010.

[3] European Centre for Disease Prevention and Control. Covid-19. https://www.ecdc.europa.eu/en/covid-19-pandemic. Accessed: 2020-04-09.

[4] Data Science for Social Good Portugal. Dados relativos à pandemia COVID-19 em portugal. https://github.com/dssg-pt/covid19pt-data. Accessed: 2020-04-09.

[5] A Maxmen. How much is coronavirus spreading under the radar? Nature, 2020. https://www.nature.com/articles/d41586-020-00760-8.

[6] Hiroshi Nishiura, Don Klinkenberg, Mick Roberts, and Johan AP Heesterbeek. Early epidemiological assessment of the virulence of emerging infectious diseases: a case study of an influenza pandemic. PLoS One, 4(8), 2009. https://journals.plos.org/plosone/article?id=10.1371/journal.pone.0006852.

[7] The @CoronaSurveys research team. @CoronaSurveys: Monitoring the incidence of COVID-19 via open surveys. https://coronasurveys.org/. Accessed: 2020-04-20.

[8] T Russel, Joel Hellewell, S Abbot, et al. Using a delay-adjusted case fatality ratio to estimate under-reporting. Available at the Centre for Mathematical Modelling of Infectious Diseases Repository, here, 2020. https://cmmid.github.io/topics/covid19/severity/global_cfr_estimates.html.

[9] Wikipedia. Dunbar’s number. https://en.wikipedia.org/wiki/Dunbar%27s_number. Accessed: 2020-04-09.

[10] Peng Yang, Chunna Ma, Weixian Shi, Shujuan Cui, Guilan Lu, Xiaomin Peng, Daitao Zhang, Yimeng Liu, Huijie Liang, Yi Zhang, et al. A serological survey of antibodies to H5, H7 and H9 avian influenza viruses amongst the duck-related workers in Beijing, China. PLoS One, 7(11), 2012. https://journals.plos.org/plosone/article?id=10.1371/journal.pone.0050770.

